# Deafferentation of Olfactory Bulb in Subjects Dying with COVID-19

**DOI:** 10.1101/2021.12.21.21268119

**Authors:** Cécilia Tremblay, Thomas G. Beach, Anthony J. Intorcia, Jessica E. Walker, Richard A. Arce, Lucia I. Sue, Courtney M. Nelson, Claryssa I. Borja, Katsuko E. Suszczewicz, Madison P. Cline, Spencer J. Hemmingsen, Sanaria H. Qiji, Marc Desforges, Geidy E. Serrano

**Author notes:** Correspondence: Thomas G. Beach, Banner Sun Health Research Institute, 10515 West Santa Fe Drive, Sun City, AZ 85351.

## Abstract

There have been clinical descriptions of diverse neurological effects in COVID-19 disease, involving up to 36% of patients. It appears likely that most of these are not caused by viral brain invasion but by systemic accompaniments of critical illness such as coagulopathy, deleteriously upregulated immune response, autoimmune mechanisms, hypoxia or multiorgan failure. Anosmia or hyposmia is present in a majority of COVID-19 patients, and there is early and severe involvement of the nasopharyngeal mucosa and olfactory epithelium. Preliminary studies by our group have found massive gene expression changes in olfactory bulb, but the magnitude of these changes are not different between subjects with detectable versus non-detectable olfactory bulb SARS-CoV-2 RNA. As spontaneous discharge of olfactory epithelial afferents dictates intra-olfactory bulb neurophysiological activity and connectivity, we hypothesized that olfactory bulb deafferentation during COVID-19 is responsible for a large fraction of our observed olfactory bulb transcriptional changes. As the olfactory marker protein (OMP-1) is a specific marker of olfactory epithelial afferents to the olfactory bulb and is severely depleted in animal model lesions of olfactory epithelium, we quantified OMP-1-immunoreactivity in the olfactory bulb of subjects dying with or without COVID-19. Additionally, we quantified olfactory bulb tyrosine hydroxylase (TH), which is often also reduced after olfactory epithelium lesions, and SNAP-25, a pan-synaptic marker. COVID-19 cases (n = 18) were generally elderly and were not significantly different in age or gender distribution from the non-COVID-19 cases (n = 28). Both COVID-19 and non-COVID-19 cases had a wide range of neuropathological diagnoses. The area occupied by OMP-1 immunoreactivity in COVID-19 cases was significantly less, about 60% of that in control cases but amongst subjects with COVID-19, there was no significant difference between OBT-SARS-CoV-2-PCR-positive and negative cases. There were no significant group differences for TH or SNAP-25, supporting a selective effect for OMP-1. We suggest that olfactory dysfunction, and some of the COVID-19-associated transcriptional changes that we have reported for the olfactory bulb and amygdala, may be due to olfactory bulb deafferentation and subsequent transsynaptic effects. Additionally, animal models of olfactory bulb deafferentation or bulbectomy indicate a possibility for widespread changes in interconnected brain regions, providing a possible substrate for diverse post-acute COVID-19 neurological sequelae.

## INTRODUCTION

The coronavirus SARS-CoV-2 is the cause of pandemic severe respiratory disease, termed COVID-19, but there have additionally been clinical descriptions of diverse neurological effects, involving up to 36% of patients ^1-10^. What remains unclear is whether these are caused by viral brain invasion or by systemic accompaniments of critical illness such as coagulopathy, deleteriously upregulated immune response, autoimmune mechanisms, hypoxia or multiorgan failure ^11^.

More than 20 published studies ^11-31^ have utilized RT-PCR methods to determine the presence of SARS-CoV-2 genomic fragments in postmortem brain, and although results have been variable, the most likely conclusion is that SARS-CoV-2 brain invasion occurs in only a smaller fraction of subjects, viral copy numbers are low, and proof of in situ viral CNS replication or protein synthesis is lacking ^13,31-35^.

Anosmia or hyposmia is present in many COVID-19 patients ^36-38^ and clinical, autopsy and animal model evidence has pointed to early and severe involvement of the nasopharyngeal mucosa and olfactory epithelium^27,31,36,39-45^. The olfactory epithelium includes olfactory sensory neurons that project directly through the perforated cribriform plate to the intracranial olfactory bulb, making this pathway a major candidate for brain entry. Of studies localizing SARS-CoV-2 RNA in multiple brain regions of deceased COVID-19 subjects, olfactory bulb has been found to be the single most-involved area^31,46^. Preliminary studies by our group have found massive gene expression changes in olfactory bulb^47^, with lesser but still marked changes in the amygdala, one of its main projection targets. The magnitude of these changes, however, are not different between subjects with detectable versus non-detectable olfactory bulb SARS-CoV-2 RNA. As spontaneous discharge of olfactory epithelial afferents dictates intra-olfactory bulb neurophysiological activity and connectivity^48-50^, we hypothesized that deafferentation during COVID-19 is responsible for the majority of our observed olfactory bulb transcriptional changes. As the olfactory marker protein (OMP-1) is a specific marker of olfactory epithelial afferents to the olfactory bulb^51-53^ and is severely depleted in animal model lesions of olfactory epithelium^48,54-57^, we quantified OMP-1-immunoreactivity in the olfactory bulb of subjects dying with or without COVID-19. Additionally, we quantified olfactory bulb tyrosine hydroxylase (TH) ^54,55,58^, which is often also reduced after olfactory epithelium lesions, and SNAP-25, a pan-synaptic marker.

## MATERIALS AND METHODS

### Human Subjects and Characterization

The COVID-19 subjects were derived from Banner Sun Health Research Institute (BSHRI) in Sun City, Arizona (n =18). Ethical approval for autopsy and subsequent research was obtained for all subjects through Institutional Review Board-approved protocols and informed consent documents (Western IRB, Puyallup, WA; protocols 1132516 and 20201852). Clinical and neuropathological results for some of the COVID-19 cases are described in our prior publications ^46,47^ and the complete set of cases used in the present study is summarized in Table 1. All COVID-19 subjects had positive clinical diagnostic test results for SARS-CoV-2 and all were considered to have died acutely or subacutely in 2020 or early 2021 as a result of COVID-19. An additional 28 cases were chosen from 2018, 2019 and 2020 non-COVID-19 BSHRI autopsies, including 13 cases with autopsy-proven non-COVID-19 pneumonia and 15 cases without autopsy evidence of pneumonia. Non-COVID-19 status was determined by autopsy occurrence prior to the COVID-19 US arrival (2018 and 2019 autopsies) or, for 2020 and 2021 cases, by a negative postmortem nasopharyngeal RT-PCR test conducted in a CLIA-approved laboratory.

**Table 1.** Case data. Olf. Bulb SCV2 = PCR-+ for SARS-CoV-2 in olfactory bulb; AcInf = Acute or subacute infarction or ischemic-hypoxic histological changes; AD = Alzheimer’s disease; ADinsuff = AD changes insufficient for diagnosis; DLB = dementia with Lewy bodies; FTLD-TDP = frontotemporal lobar degeneration with TDP-43 proteinopathy; HS = hippocampal sclerosis; LATE = Limbic age-related TDP-43 proteinopathy; LBinsuff = Lewy body disease insufficient for diagnosis of PD or DLB; PD = Parkinson’s disease; PART = primary age-related tauopathy; PSP = microscopic changes consistent with progressive supranuclear palsy; VaD = vascular dementia. For all subjects, exact age is not specified due to privacy regulations. N/A = not available or not done. Yellow = Major neuropathological diagnosis.

| Case | Age | Sex | COVID-19 | Olf. Bulb SCV2 | Non-COVID-19 Pneumonia | Neuropathologically-Diagnosed Conditions |
| --- | --- | --- | --- | --- | --- | --- |
| B1 | 90s | M | + | - | - | AD; VaD; HS; PSP; TDP-43 (LATE) |
| B2 | 70s | F | + | - | - | PD; AD; DBS |
| B3 | 70s | M | + | - | - | PART; AcInf |
| B4 | 80s | M | + | + | - | ADinsuff |
| B5 | 90s | M | + | + | - | ADinsuff |
| B6 | 70s | M | + | - | - | AD; TDP-43 (LATE) |
| B7 | 70s | F | + | + | - | ADinsuff |
| B8 | 80s | M | + | - | - | AD; MSA; TDP-43 (LATE); AcInf |
| B9 | 70s | M | + | + | - | AD insuff |
| B10 | 60s | F | + | - | - | PART |
| B11 | 90s | F | + | - | - | AD; DLB |
| B12 | 80s | F | + | + | - | AD; TDP-43 (LATE) |
| B13 | 90s | M | + | - | - | PSP; ADinsuff |
| B14 | 80s | F | + | - | - | VaD |
| B15 | 90s | M | + | - | - | AD; DLB |
| B16 | 90s | M | + | - | - | AD; TDP-43 (LATE) |
| B17 | 80s | F | + | - | - | AD; TDP-43 (LATE) |
| B18 | 70s | M | + | - | - | PART |
| C1 | 70s | M | - | N/A | - | PART |
| C2 | 70s | F | - | N/A | - | ADinsuff |
| C3 | 90s | M | - | N/A | + | AD-insuff; LBinsuff |
| C4 | 80s | F | - | N/A | - | AD-insuff; TDP-43 (LATE) |
| C5 | 90s | F | - | N/A | - | AD-insuff; LATE |
| C6 | 100s | M | - | N/A | + | PART; LATE |
| C7 | 70s | F | - | N/A | - | ADinsuff |
| C8 | 80s | M | - | N/A | + | AD; AcInf |
| C9 | 70s | M | - | N/A | + | PART; AcInf |
| C10 | 70s | M | - | N/A | + | AD; PD |
| C11 | 70s | M | - | N/A | + | AD; PD |
| C12 | 80s | F | - | N/A | - | AD; VaD; LBinsuff |
| C13 | 80s | M | - | N/A | + | AD; TDP-43 (LATE); LBinsuff |
| C14 | 80s | M | - | N/A | + | PD; PART; TDP-43 (LATE) |
| C15 | 90s | M | - | N/A | - | AD; VaD; AcInf; TDP-43 (LATE) |
| C16 | 90s | F | - | N/A | - | AD; TDP-43 (LATE) |
| C17 | 90s | F | - | N/A | - | AD; PSP; HS; FTLTDP; VaD |
| C18 | 80s | M | - | N/A | + | AD; DLB; VaD; PSP; TDP-43 (LATE) |
| C19 | 90s | M | - | N/A | - | ADinsuff; LBinsuff; AcInf |
| C20 | 90s | F | - | N/A | + | AD; TDP-43 (LATE) |
| C21 | 90s | M | - | N/A | + | ADinsuff; TDP-43 (LATE); AcInf |
| C22 | 90s | F | - | N/A | + | AD |
| C23 | 90s | M | - | N/A | + | AD; LBinsuff; TDP-43 (LATE); PSP |
| C24 | 100s | F | - | N/A | + | VaD; TDP-43 (LATE) |
| C25 | 80s | M | - | N/A | + | AD; FTLTDP; HS; PSP; LBinsuff |
| C26 | 70s | M | - | N/A | - | PART |
| C27 | 80s | M | - | N/A | - | MSA; ADinsuff |
| C28 | 80s | M | - | N/A | - | ADinsuff; TDP-43 (LATE) |

Published diagnostic clinicopathological consensus criteria for age-related and neurodegenerative brain disease ^59-71^ were used when applicable, incorporating research clinical assessment results as well as pertinent private medical history. The histological sampling and staining incorporated the protocols recommended by the National Institute on Aging and Alzheimer’s Association (NIA-AA) ^69-71^.

Immunohistochemical staining for OMP-1, TH and SNAP-25 was done with commercially obtained antibodies (OMP-1 Novus Biologicals NB110-74751; TH Sigma Aldrich T2928; SNAP-25 Sigma-Aldrich S9684) on formalin-fixed, paraffin-embedded sections of olfactory bulb. Epitope exposure methods used included 20 minutes in boiling 0.1 M sodium citrate for OMP-1 and TH, and 20 minutes in 80% formic acid for SNAP-25. Primary antibody concentrations used were 1;2,000 for OMP-1, 1:3,000 for TH and 1:5,000 for SNAP-25. Control sections omitted the primary antibodies. Four images per case were captured from slides stained with each antibody. The final magnifications used were 200x for OMP-1 and 400x for TH and SNAP25. Image capture and analyses were done while blinded to subject clinical or autopsy information.

The area occupied by stained tissue elements within the olfactory bulb nerve fiber layer and adjacent glomerular layer was determined with digital image analysis (Image J software with image processing and analysis in Java: https://imagej.nih.gov/ij/). The means of the areas occupied by staining in the 4 images were used for statistical analyses, performed using SPSS software (IBM SPSS Statistics 23.0). Methods included unpaired, two-way t-tests for continuous variables, as well as analysis of variance with post-hoc Bonferroni-corrected pairwise significance testing. Fisher Exact tests were used to test for proportional differences and linear regression for relationships between continuous variables. The probability level was set at p < 0.05.

## RESULTS

Table 1 shows basic data for the cases studied. COVID-19 cases did not significantly differ in age from the non-COVID-19 cases (82.3, SD 8.9 vs 86.2, SD 9.2, respectively). Males made up 11/18 and 18/28 of the COVID-19 and non-COVID-19 cases, respectively (ns). Both COVID-19 and non-COVID-19 cases had a wide range of neuropathological diagnoses, consistent with their age and derivation from BSHRI, a research center devoted to the study of aging and neurodegenerative disease.

Figures 1 and 2 show photomicrographs and quantification results, respectively, for OMP-1, TH and SNAP-25 immunoreactivity in COVID-19 and non-COVID-19 control cases. As expected from previous reports, positive OMP-1 staining was restricted to the peripheral nerve fiber and glomerular cell layers while TH and especially SNAP-25 were more diffusely distributed throughout the bulb. The area occupied by OMP-1 immunoreactivity in COVID-19 cases was significantly less, about 60% of that in control cases; t(43) = -2.908; p= 0.006 (Figure 2a). Analysis of variance showed an overall difference in OMP-1 immunoreactivity between the groups when controls were subdivided by the presence or absence of non-COVID-19 pneumonia; F (2,42) = 4.176; p = 0.022 (Figure 2a). Pairwise post hoc significance testing showed significant differences between the COVID-19 group and the controls without pneumonia (p = 0.048) but no differences between the COVID-19 group and the controls with pneumonia. There was no statistical difference between controls with and without pneumonia.

**Figure 1:**
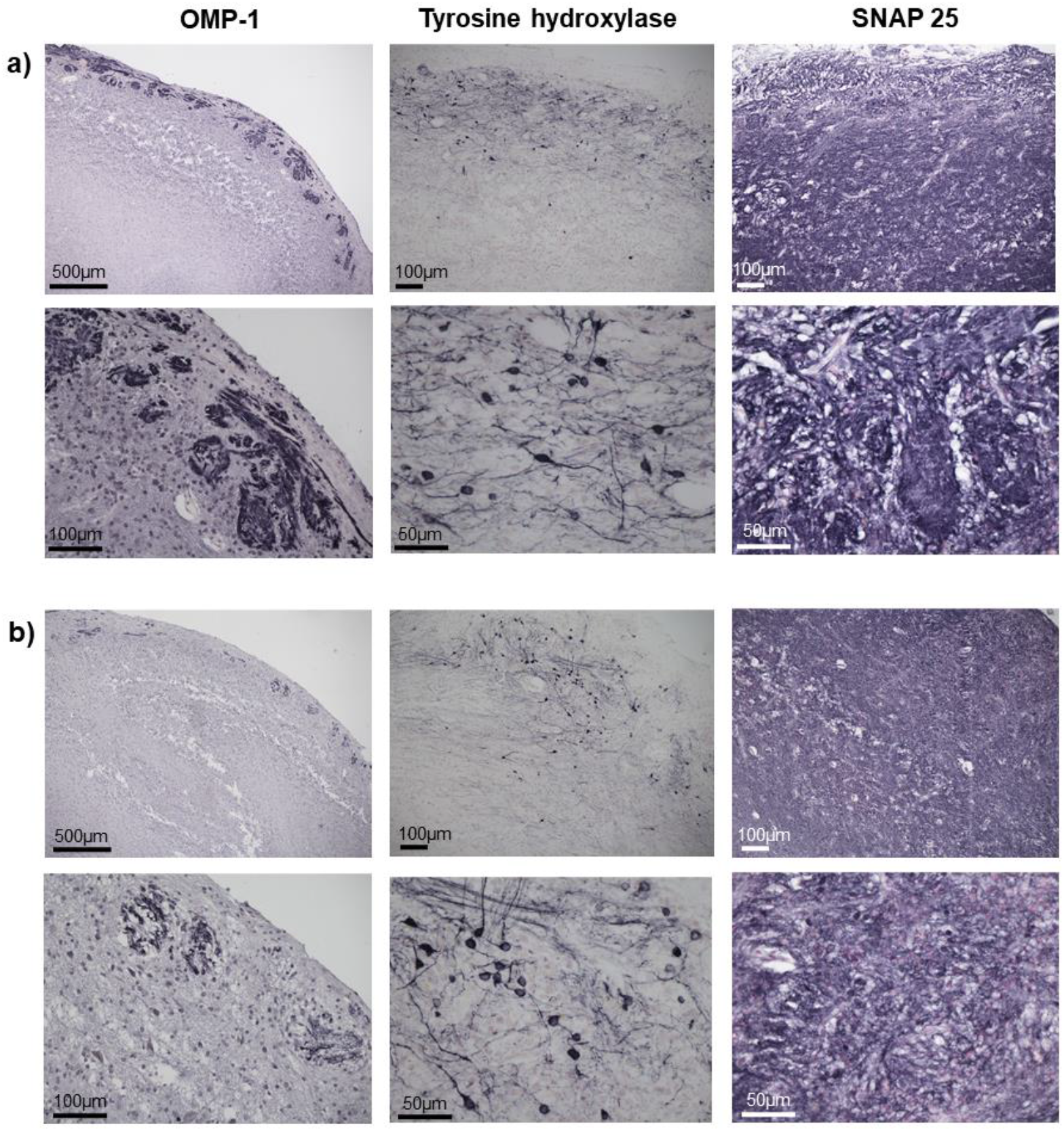
Photomicrographs representative of OMP-1, TH and SNAP-25 immunoreactivity in a) non-COVID-19 control cases and b) in COVID-19 cases at low and high magnifications, in sections of olfactory bulb. The second row for each set shows the magnification used for image analysis. Immunoreactivity for OMP-1 (dark purple) consists solely of fibers and puncta, consistent with axons and presynaptic terminals, and is restricted to the nerve fiber layer and glomerular layer, as expected from the known termination of nasopharyngeal olfactory sensory afferents. Staining for TH and particularly SNAP-25 (both are dark purple) is distributed more widely. Neuronal cell bodies and dendrites are apparent in the TH-stained sections while only fibers and puncta, consistent with axons, dendrites and synaptic terminals, are seen in the SNAP-25-stained sections.

**Figure 2:**
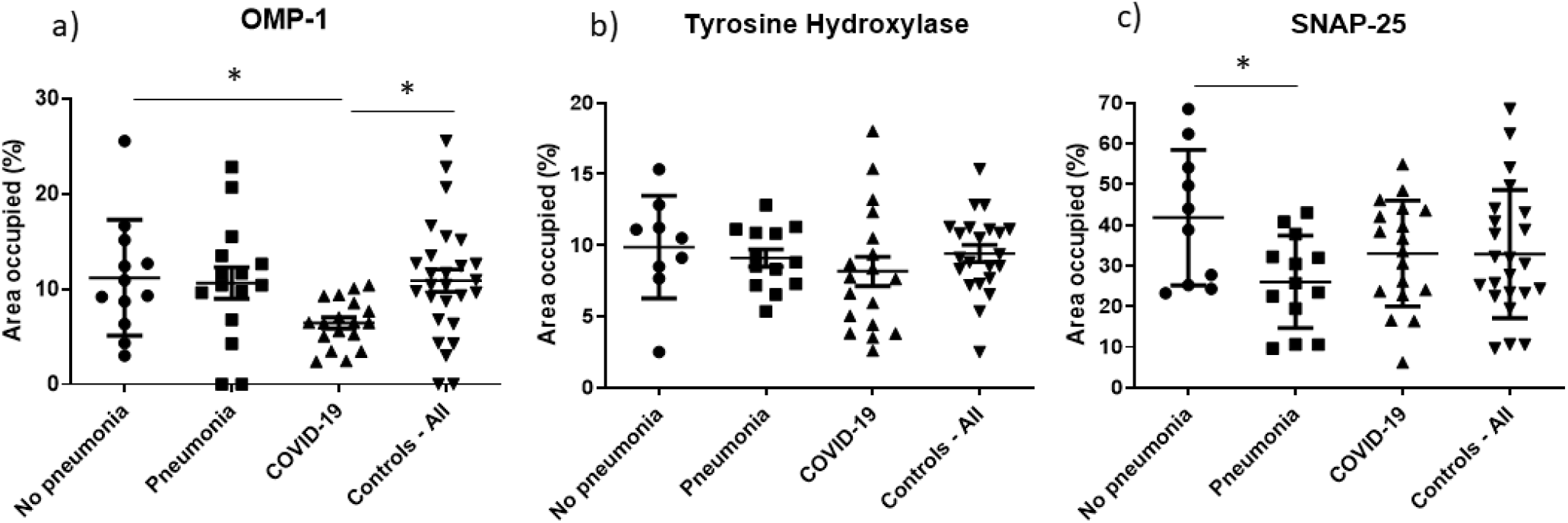
Quantification results for OMP-1, TH and SNAP-25 immunoreactivity for each group. All controls include both controls with and without pneumonia. Means and standard deviations of the means are represented. * = p < 0.05 (see text for precise p values).

Analysis of variance showed no significant group differences for TH (Figure 2b) while for SNAP-25 (Figure 2c) the group difference was significant; F (2,38) = 3.878; p = 0.029. Significantly less SNAP-25 staining was present in controls with pneumonia as compared to controls without pneumonia (p = 0.025) but there were no significant differences between COVID-19 cases and all controls, or between COVID-19 cases and controls subdivided by presence or absence of pneumonia.

As COVID-19 cases differed with respect to the presence or absence of olfactory bulb SARS-CoV-2 RNA, as detected by RT-PCR, we compared immunoreactivity for the 3 antibodies in the PCR-positive and PCR-negative cases versus all controls. For OMP-1, analysis of variance showed a significant difference between groups; F (2,42) = 4.251; p = 0.02. Post hoc significance testing showed a significant difference between OBT-PCR-negative COVID-19 cases and controls (p = 0.027) but no significant difference between COVID-19 OBT-PCR-positive and PCR-negative cases. Analysis of variance of the same groups for TH or SNAP-25 staining showed no significant group differences (p = 0.239 and 0.714, respectively).

Comparing COVID-19 cases and controls for the presence or absence of a major, neuropathologically-diagnosed neurodegenerative or cerebrovascular disease, analysis of variance showed no significant difference between groups, with any of the 3 antibodies.

## DISCUSSION

COVID-19 is primarily a respiratory disease but with up to 36% of patients showing neurological effects ^1-10^, there is enormous potential for this additional morbidity and so it is of high importance that the mechanisms of CNS impairment be understood. At present the relative contributions of direct viral brain invasion versus indirect factors are unclear. The most unequivocal direct viral manifestation is acute encephalitis, with a reported mortality rate of 13%, but this has been estimated to occur in only 0.2% of all cases ^72^; furthermore, this is likely an overestimate as virtually all diagnoses have been made radiologically and could not have distinguished between autoimmune mechanisms versus direct viral brain infection. Systemic accompaniments of critical illness, including coagulopathy, aberrant immune responses, hypoxia and multi-organ failure are therefore more likely to account for most of the reported COVID-19-associated neurological syndromes ^11^.

Postmortem brain tissue studies support this likelihood, with more than 20 published studies ^11-31^ using RT-PCR in general agreement that SARS-CoV-2 brain invasion occurs in only a smaller fraction of subjects, and in these the viral copy numbers are often low ^13,31-35^. In our own comprehensive brain mapping of SARS-CoV-2 RNA in 320 brain regions of 20 deceased COVID-19 subjects, the olfactory bulb has been found to be the single most-involved area^46,47^ but still was positive in only 8 of 20 cases. More than 5,000 gene expression changes were present in the olfactory bulb in COVID-19 cases as compared to control autopsies, but there were no differences between COVID-19 cases with positive versus negative RT-PCR results ^47^. Fewer gene expression changes, but still more than 1,000, were found in the amygdala, a main projection target of the olfactory bulb, despite only 2/20 cases being RT-PCR positive there. Pathway analysis for the amygdala indicated enrichment and downregulation, respectively, of immune signaling and neuronal/synaptic networks while in olfactory bulb the top upregulated pathway was related to olfactory signaling.

As SARS-CoV-2 invariably infects the nasopharyngeal mucosa and associated olfactory epithelium ^27,31,36,39-45^, frequently resulting in hyposmia ^36-38^, we hypothesized that a large fraction of COVID-19-related olfactory bulb transcriptional changes may be due to its deafferentation during COVID-19, as spontaneous discharge of olfactory epithelial afferents is a dominant regulator of olfactory bulb neurophysiological activity and connectivity^48-50^.

In this study we report that the olfactory marker protein (OMP-1), a specific marker of olfactory epithelial afferents to the olfactory bulb^51-53^, is significantly depleted in the olfactory bulb of subjects dying with COVID-This parallels what has been reported in animal model lesions of olfactory epithelium^48,54-57^. Unlike in some animal models ^54,55,58^, we did not find that olfactory bulb tyrosine hydroxylase is significantly depleted in the COVID-19 subjects, but this may be dependent on the time elapsed from deafferentation. There was no depletion of SNAP-25, a protein found in most or all neuronal synapses, supporting a selective effect of COVID-19 on olfactory epithelial axons. Neither gene expression changes or OMP-1 immunoreactivity were significantly different in COVID-19 olfactory bulbs with or without positive RT-PCR for SARS-CoV2 RNA, reinforcing an emerging recognition that CNS alterations in COVID-19 may, in the majority of subjects, occur independently of direct viral action.

We suggest that at least some of the COVID-19-associated transcriptional changes that we have reported for the amygdala may be due to transsynaptic effects initiated by olfactory bulb deafferentation, as the amygdala and other brain regions are directly innervated by the olfactory bulb. Olfactory stimulation activates neurons of the amygdala ^73^, while olfactory bulbectomy in mice has been reported to cause piriform cortex reactions including activation of interneurons, apoptosis of pyramidal neurons and downregulation of regulatory pathways ^74-79^. A UK Biobank imaging study of 401 subjects before and after SARS-CoV-2 infection ^80^ found markers of tissue damage were primarily in olfactory-related brain regions including the medial and lateral orbitofrontal cortex, the anterior insula, the anterior cingulate cortex and the amygdala. Metabolic imaging in patients with “long COVID” found hypometabolism in the amygdala, hippocampus, and bilateral rectal and orbital gyri ^81^, similarly to the decreased global cerebral glucose utilization reported after olfactory bulbectomy in an animal model ^82^. A recent study has reported evidence that SARS-CoV-2 infection of supporting olfactory epithelium sustentacular cells may be the cause of olfactory sensory dysfunction in COVID-19 ^83^.

Transsynaptic changes following olfactory deafferentation may have diverse behavioral effects. Olfactory bulbectomy is the basis of a rat model of depression ^84-86^ associated with inflammatory and intermediate early gene expression in the amygdala ^87,88^, and olfactory bulbectomy has also been associated with memory and cognitive deficits thought to be related to basal forebrain cholinergic effects ^89-94^, providing a possible substrate for diverse post-acute COVID-19 neurological sequelae.

In conclusion, we report a significant depletion, in olfactory bulb of subjects dying with COVID-19, of OMP-1, a specific marker of primary olfactory sensory input. This may be a structural correlate of the clinical hyposmia reported for many subjects with COVID-19. Associated transsynaptic effects may result in more widespread changes in interconnected brain regions. Limitations of our study include relatively small subject numbers, especially for subgroups, and inclusion of only COVID-19 subjects with severe illness.

## Data Availability

All data produced in the present study are available upon reasonable request to the authors.

## ACKNOWLEDGEMENTS

This project was supported by a Covid-19 Supplement to a National Institute on Aging grant, (3P30AG019610-20S1), submitted in response to a Notice of Special Interest (NOSI) issued by the National Institute on Aging (NOT-AG-20-022), to highlight the urgent need for research on Coronavirus Disease 2019….

